# New Serum Potassium Cut-Off Point for Improving Primary Aldosteronism Screening

**DOI:** 10.64898/2026.06.30.26356983

**Authors:** Hailin Li, Fen Zhou, Hongchao Zhao, Wenli huang, Haibo Wang, Shuo Wang

## Abstract

This study enrolled 152 hypertensive patients with an ARR > 3.7 to assess the relationship between the traditional hypokalemia cutoff (3.5 mmol/L) and primary aldosteronism (PA) screening, and to establish a new cutoff. Under the traditional cutoff, only 35.7% of PA patients presented with hypokalemia. ROC curve analysis identified a new cutoff of 4.22 mmol/L, which increased sensitivity from 35.7% to 77.5%, with a specificity of 91.1% and an AUC of 0.897. The findings indicate that the traditional cutoff is insufficiently sensitive, while the new cutoff markedly improves screening sensitivity and facilitates early detection of PA.

## 1. Introduction

Primary aldosteronism (PA) is the most prevalent form of secondary hypertension, among patients with grade 1, 2, and 3 hypertension the prevalence of primary aldosteronism was 1.99%, 8.02% and 13.2% respectively^1^ and it was even higher in patients with refractory hypertension approximately 17% - 23% ^2^. Studies have found that excessive aldosterone is an important risk factor for myocardial hypertrophy, heart failure, and renal function impairment. Compared with patients with primary hypertension, patients with primary aldosteronism have more severe damage to hypertension target organs such as the heart and kidneys. Therefore, early diagnosis and treatment are of crucial importance.

The ARR is the most widely used screening test for PA, but its specificity is limited (estimated 30– 50%) ^3,4^, leading to a high rate of false-positive results and unnecessary confirmatory testing. Conversely, the conventional hypokalemia threshold of 3.5 mmol/L is highly specific (approaching 100%) but insensitive, identifying fewer than 30% of PA patients^5^. This means that the majority of PA patients (≥70%) have serum potassium levels within the normal range, often in the low-normal range, and thus fail to trigger clinical suspicion when hypokalemia is used as the sole trigger^4-6^. Therefore, combining ARR (sensitive but non-specific) with a refined potassium threshold (specific but currently under-used) could offer a balanced approach. In this study, we evaluated whether optimizing the potassium cut-off in ARR-positive patients improves the diagnostic yield. Specifically, we hypothesized that in hypertensive patients with an elevated ARR, refining the potassium threshold could improve PA detection. In this study, we determined a new serum potassium cut-off value (4.22 mmol/L) among ARR-positive individuals, aiming to enhance screening sensitivity while maintaining acceptable specificity, and thereby increase the overall yield of PA diagnosis. The present study aimed to: (1) determine the prevalence of hypokalemia (serum potassium <3.5 mmol/L) among patients with confirmed PA; (2) characterize the distribution of serum potassium across PA, IHA, and PH groups; and (3) establish a new serum potassium cut-off point in ARR positive patients for PA screening through systematic ROC curve analysis, with the goal of improving case detection sensitivity while maintaining acceptable specificity.

## 2. Materials and Methods

### 2.1. Study Design and Subjects

This cross-sectional study was conducted at the Department of Endocrinology, Renmin Hospital of Wuhan University, between January 2021 and December 2025. A total of 152 consecutive patients with hypertension were enrolled and classified into three groups: APA(n=72), IHA (n=40), and PH (n=40). The study was approved by the Ethics Committee of Renmin Hospital of Wuhan University, and written informed consent was obtained from all participants prior to enrollment.

Inclusion criteria: (1) Age 18–75 years; (2) Confirmed diagnosis of hypertension (systolic blood pressure ≥140 mmHg and/or diastolic blood pressure ≥90 mmHg, or current use of antihypertensive medications); (3) Completion of standardized hormonal evaluation including ARR screening, confirmatory testing, and serum potassium measurement.

Exclusion criteria: (1) Use of medications that significantly affect serum potassium or aldosterone/renin measurements and could not be discontinued for at least 4 weeks prior to testing (e.g., spironolactone, eplerenone, potassium-sparing diuretics, potassium supplements); (2) Renal insufficiency (estimated glomerular filtration rate <30 mL/min/1.73 m^2^); (3) Known causes of secondary hypertension other than PA (e.g., pheochromocytoma, Cushing’s syndrome, renal artery stenosis); (4) Pregnancy; (5) Incomplete clinical or laboratory data.

### 2.2. Diagnostic Criteria and Grouping

PA diagnosis was established according to the Clinical Practice Guidelines^7^. All patients with an elevated ARR (>3.7) underwent confirmatory testing with the saline infusion test (SIT). PA was confirmed if post-SIT plasma aldosterone concentration (PAC) remained ≥10 ng/dL. Adrenal venous sampling (AVS) was performed in confirmed PA patients to differentiate APA from IHA.

PH diagnosis was established in hypertensive patients with an elevated ARR (>3.7), SIT-negative and no clinical or imaging evidence of secondary hypertension. Furthermore, all the patients’ low-dose dexamethasone suppression test were negative.

### 2.3. Biochemical Evaluation

All patients underwent standardized biochemical evaluation under controlled conditions. Antihypertensive medications affecting the renin-angiotensin-aldosterone system (RAAS) were discontinued or substituted with non-interfering agents for at least 2 weeks prior to testing. Spironolactone and eplerenone were discontinued for at least 6 weeks. Patients were instructed to maintain their usual dietary sodium and potassium intake and to avoid potassium supplementation unless clinically indicated.

Blood samples for PAC, DRC, and serum potassium were collected in the morning after the patient had been in an upright/seated position for at least 2 hours. PAC was measured using a chemiluminescent immunoassay. DRC was measured by radioimmunoassay. ARR was calculated as PAC (ng/dL) divided by DRC (ng/L). Serum potassium was measured using an automated ion-selective electrode analyzer. Conventional hypokalemia was defined as serum potassium <3.5 mmol/L.

### 2.4. Data Collection

Demographic and clinical data were collected, including age, sex, duration of hypertension, systolic and diastolic blood pressure (SBP, DBP), number of antihypertensive medications, serum sodium, serum potassium, PAC, DRC, ARR, and adrenal imaging findings (computed tomography).

### 2.5. Statistical Analysis

Continuous variables were expressed as mean ± standard deviation (SD) or median (interquartile range, IQR), depending on the distribution assessed by the Shapiro-Wilk test. Categorical variables were expressed as frequencies and percentages. Comparisons among the three groups were performed using one-way analysis of variance (ANOVA) or the Kruskal-Wallis test for continuous variables, and the chi-square test or Fisher’s exact test for categorical variables. Post hoc pairwise comparisons were conducted with Bonferroni correction.

The primary analysis was the determination of the optimal serum potassium cut-off point for PA screening using receiver operating characteristic (ROC) curve analysis. The area under the curve (AUC) with 95% confidence interval (CI) was calculated. The Youden index (J = sensitivity + specificity − 1) was used to identify the optimal balance between sensitivity and specificity. All statistical analyses were performed using IBM SPSS Statistics version 29.0 (IBM Corp., Armonk, NY, USA). A two-sided P value <0.05 was considered statistically significant.

## 3. Results

### 3.1. Baseline Characteristics

The baseline demographic and clinical characteristics of the 152 patients are summarized in Table 1. The PA group (n=112) included 72 patients with APA and 40 with IHA. There were no significant differences in age and sex distribution the three groups. The PA group had a significantly longer duration of hypertension compared with the PH group (9.12± 6.65 vs. 3.25± 2.99 years, P<0.001) and required more antihypertensive medications (2.167 ± 0.55 vs. 1.23± 0.42, P<0.001). Serum potassium was significantly lower (3.62 ± 0.52 vs. 4.56 ± 0.54 mmol/L, P<0.001). The PA group had higher PAC, lower DRC, and higher ARR compared with PH group (all P<0.001).

**Table 1.** Baseline characteristics of study participants.

| Characteristics | PA |  |  | PH (n=40) | P |
| --- | --- | --- | --- | --- | --- |
|  | APA (n=72) | IHA(n=40) | P |  |  |
| Age (year) | 59.14±9.97 | 57.45±9.44 | 0.383 | 55.83±8.69 | 0.124 |
| Gender (male/female) | 40/32 | 19/21 | 0.436 | 24/16 | 0.464 |
| Duration of HT (years) | 9.08±6.71 | 9.20±6.63 | 0.93 | 3.25±2.99 | <0.001 |
| SBP (mmHg) | 151.94±19.45 | 139.63±21.33 | 0.01 | 163.50±11.86 | <0.001 |
| DBP (mmHg) | 89.34±11.23 | 83.98±13.66 | 0.013 | 91.6±4.56 | 0.039 |
| No. of antihypertensive meds | 2.32±0.47 | 1.9±0.59 | <0.001 | 1.225±0.42 | <0.001 |
| Aldosterone (ng/dL) | 30.08<br>(20.91-47.97) | 17.25<br>(13.4-23.32) | <0.001 | 15.22<br>(11.72-17.29) | <0.001 |
| direct renin concentration (ng/L) | 1.17<br>(0.53-2.29) | 2.51<br>(0.83-4.06) | 0.21 | 3.31<br>(2.89-4.29) | <0.001 |
| ARR | 24.15<br>(11.85-100) | 6.33<br>(4.32-13.09) | <0.001 | 4.00<br>(3.89-4.14) | <0.001 |
| Serum K<sup>+</sup> (mmol/L) | 3.42±0.44 | 3.97±0.48 | <0.001 | 4.57±0.54 | <0.001 |
| eGFR | 95.96±16.82 | 94.72±14.63 | 0.371 | 95.20±11.73 | 0.944 |

Data are presented as mean ± SD, n (%), or \*\*median (IQR). \* $P < 0.05$ . Abbreviations: PA, primary aldosteronism; APA, aldosterone-producing adenoma; IHA bilateral idiopathic hyperaldosteronism; PH, primary hypertension; HT, hypertension; SBP, systolic blood pressure; DBP, diastolic blood pressure; PAC, plasma aldosterone concentration; DRC, direct renin concentration; ARR, aldosterone-to-renin ratio. If DRC is not detected, it is defaulted to 0 and ARR is defaulted to 100.

### 3.2. Distribution of Serum Potassium Across Groups

The distribution of serum potassium across the three groups is presented in Table 2. The PA group had a markedly left-shifted potassium distribution compared with the PH group. In the PA group, 35.7% (40/112) of patients had serum potassium <3.5 mmol/L, 25.9% (29/112) had levels between 3.5 and 4.0 mmol/L, 44.6% (50/112) between 3.5 and 34.0 mmol/L. In contrast, no patients with PH had potassium <3.5 mmol/L, 7% (7/40) had levels between 3.5 and 4.0 mmol/L, and 82.5% (33/40) had levels ≥4.0 mmol/L. The IHA group showed an intermediate distribution: 15% (6/40) <3.5 mmol/L.

**Table 2.** Distribution of serum potassium across diagnostic groups.

| Serum K<sup>+</sup> | PA |  | P | PH<br>(n=40) , n<br>(%) | P |
| --- | --- | --- | --- | --- | --- |
|  | APA<br>(n=72) , n<br>(%) | IHA<br>(n=40),<br>n (%) |  |  |  |
| <3.0mmol/L | 13 (18.1) | 1 (2.5) | 0.017 | 0 (0) | 0.002 |
| 3.0–3.5mmol/L | 21 (29.1) | 5 (12.5) | 0.045 | 0 (0) | <0.01 |
| 3.5-4.0 mmol/L | 34 (47.2) | 16 (40) | 0.461 | 7 (17.5) | 0.007 |
| ≥4.0mmol/L | 4 (5.5) | 18 (45) | <0.01 | 33 (82.5) | <0.01 |

Abbreviations: PA, primary aldosteronism; APA, aldosterone-producing adenoma; IHA bilateral idiopathic hyperaldosteronism; PH, primary hypertension.

Notably, among the 112 PA patients, 80.4% (90/112) had serum potassium below 4.0 mmol/L—a proportion far exceeding the 35.7% identified by the conventional threshold of 3.5 mmol/L. This finding underscores the fact that the majority of PA patients reside in the “low-normal” potassium range rather than in the frankly hypokalemic range.

### 3.3. Determination of New Serum Potassium Cut-Off Point

ROC curve analysis was performed to evaluate the diagnostic performance of serum potassium for distinguishing PA from PH. The AUC for serum potassium was 0.897(95% CI: 0.844–0.949, P<0.001) (Figure 1). Using the Youden index, the optimal serum potassium cut-off was determined to be 4.22 mmol/L. At this threshold, the sensitivity improved dramatically from 35.7% to 77.5%, while specificity decreased from 100% to 91.1% (Table3). ROC curve analysis was performed to evaluate the diagnostic performance of serum potassium for distinguishing APA from PH. The AUC for serum potassium was 0.958 (95% CI:0.924–0.991, P<0.001) (Figure 2 and Table4). ROC curve analysis was performed to evaluate the diagnostic performance of serum potassium for distinguishing IHA from PH. The AUC for serum potassium was 0.795(95% CI: 0.706–0.883, P<0.001) (Figure 3 and Table5).

**Figure 1.**
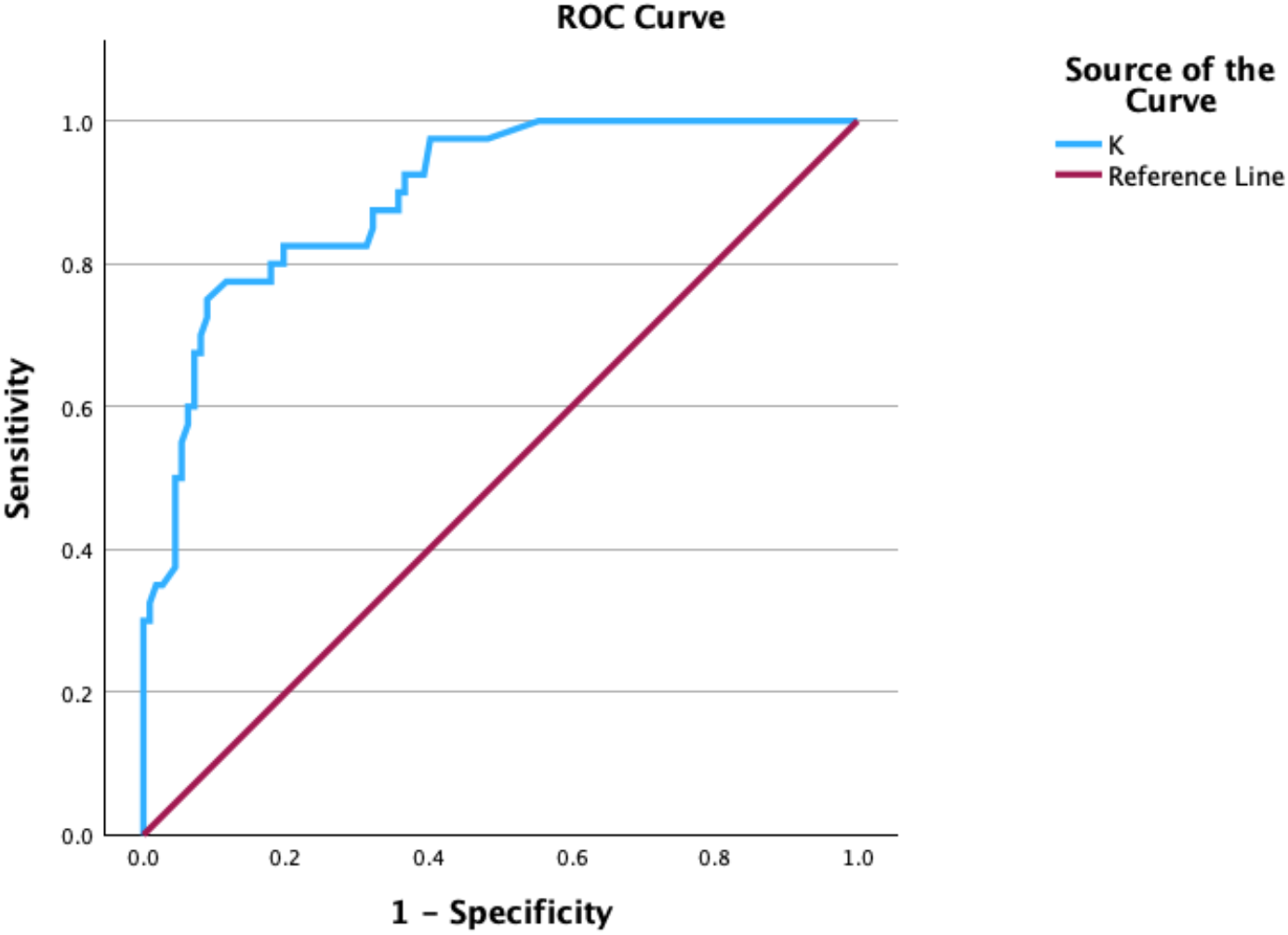
ROC curve for PA diagnosis using Serum Potassium

**Figure 2.**
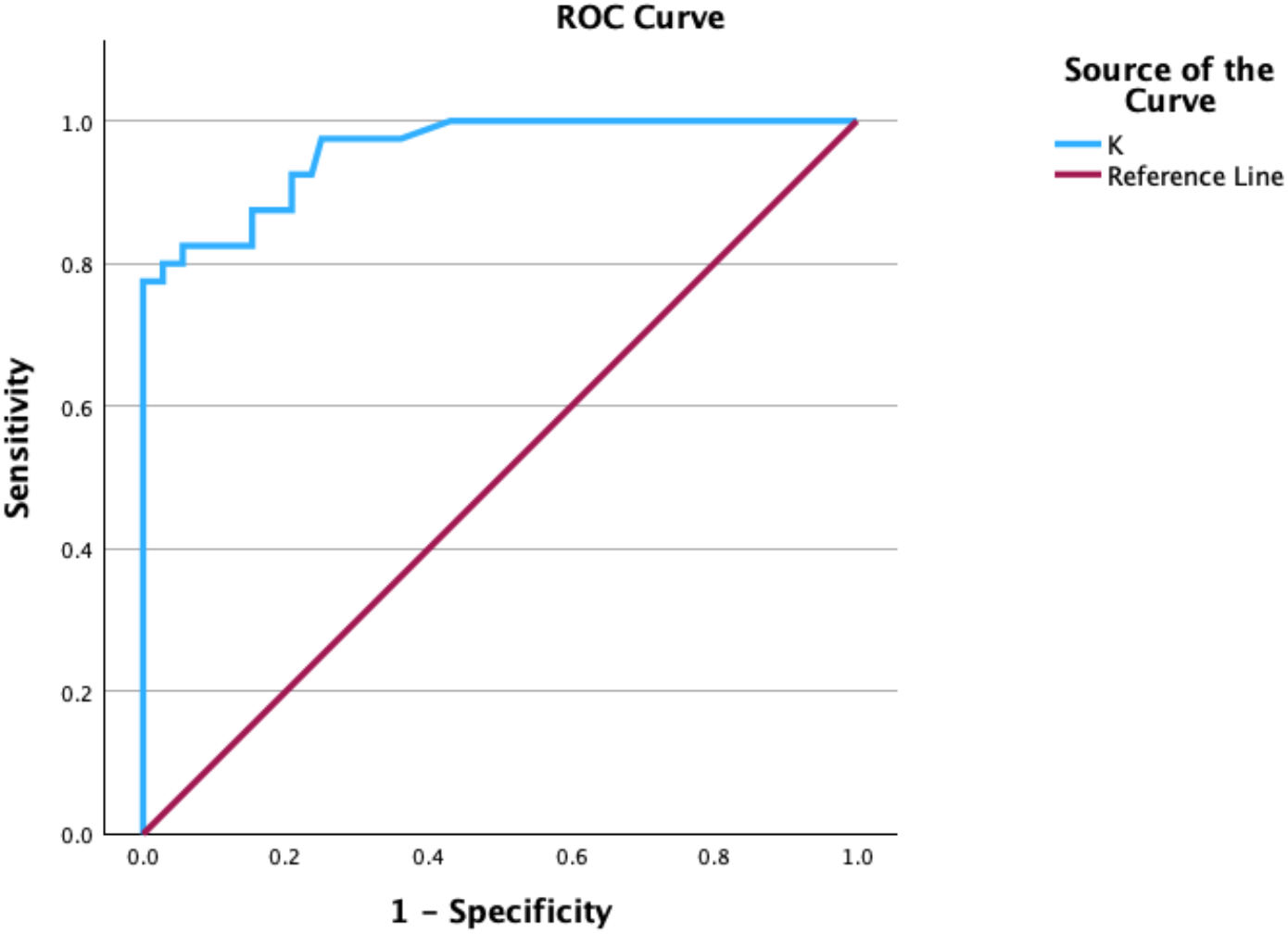
ROC curve for APA diagnosis using Serum Potassium

**Figure 3.**
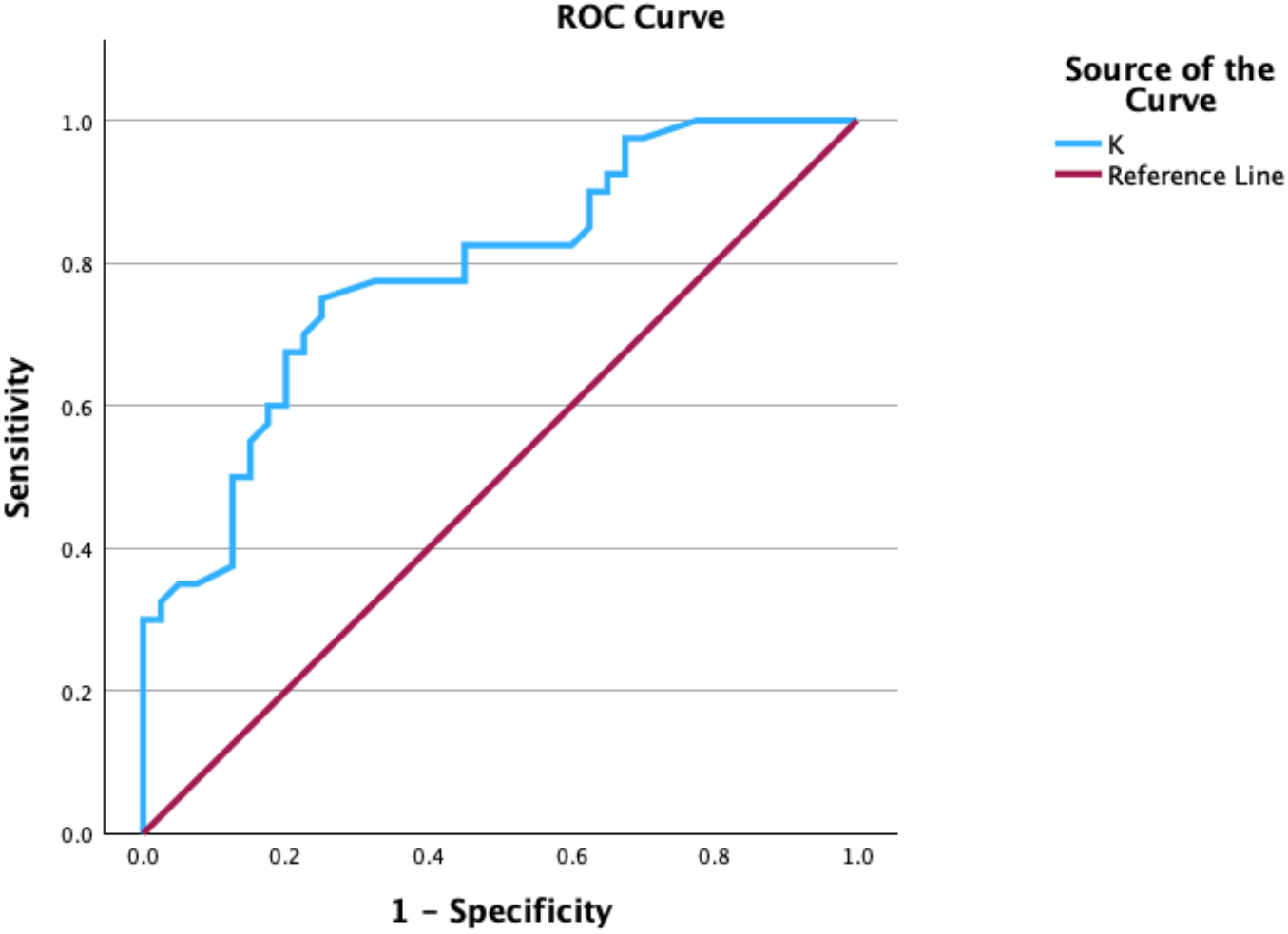
ROC curve for APA diagnosis using Serum Potassium

**Table 3.** Diagnostic performance of serum potassium cut-off points for PA detection.

| Variable | Cut-off (mmol/L) | Sen. (%) | Spec. (%) | AUC |
| --- | --- | --- | --- | --- |
| K<sup>+</sup> (conventional) | <3.5 | 35.7 | 100 | 0.357 |
| K<sup>+</sup> (new) | <4.22 | 77.5 | 91.1 | 0.897 |

Abbreviations: K<sup>+</sup>, serum potassium; Sen., sensitivity; Spec., specificity; AUC, area under the curve.

**Table 4.** Diagnostic performance of serum potassium cut-off points for APA detection.

| Variable | Cut-off (mmol/L) | Sen. (%) | Spec. (%) | AUC |
| --- | --- | --- | --- | --- |
| K<sup>+</sup> (conventional) | <3.5 | 47.2 | 100 | 0.472 |
| K<sup>+</sup> (new) | <3.98 | 94.4 | 82.5 | 0.958 |

Abbreviations: K<sup>+</sup>, serum potassium; Sen., sensitivity; Spec., specificity; AUC, area under the curve.

**Table 5.** Diagnostic performance of serum potassium cut-off points for IHA detection.

| Variable | Cut-off (mmol/L) | Sen. (%) | Spec. (%) | AUC |
| --- | --- | --- | --- | --- |
| K<sup>+</sup> (conventional) | <3.5 | 15 | 100 | 0.15 |
| K<sup>+</sup> (new) | <4.22 | 83.5 | 94.4 | 0.787 |

Abbreviations: K<sup>+</sup>, serum potassium; Sen., sensitivity; Spec., specificity; AUC, area under the curve.

### 3.4. Prevalence of Hypokalemia Using Conventional Cut-Off

Using the conventional hypokalemia cut-off of <3.5 mmol/L, only 40 of 112 PA patients (35.7%) were identified as hypokalemic (Table 6). The sensitivity of this threshold for PA detection was unacceptably low at 35.7%, despite a specificity of 100% (70/70). Among PA subtypes, hypokalemia was more prevalent in APA patients (47.2%, 34/72) compared with IHA patients (15%, 6/40) (P<0.01).

**Table 6.**
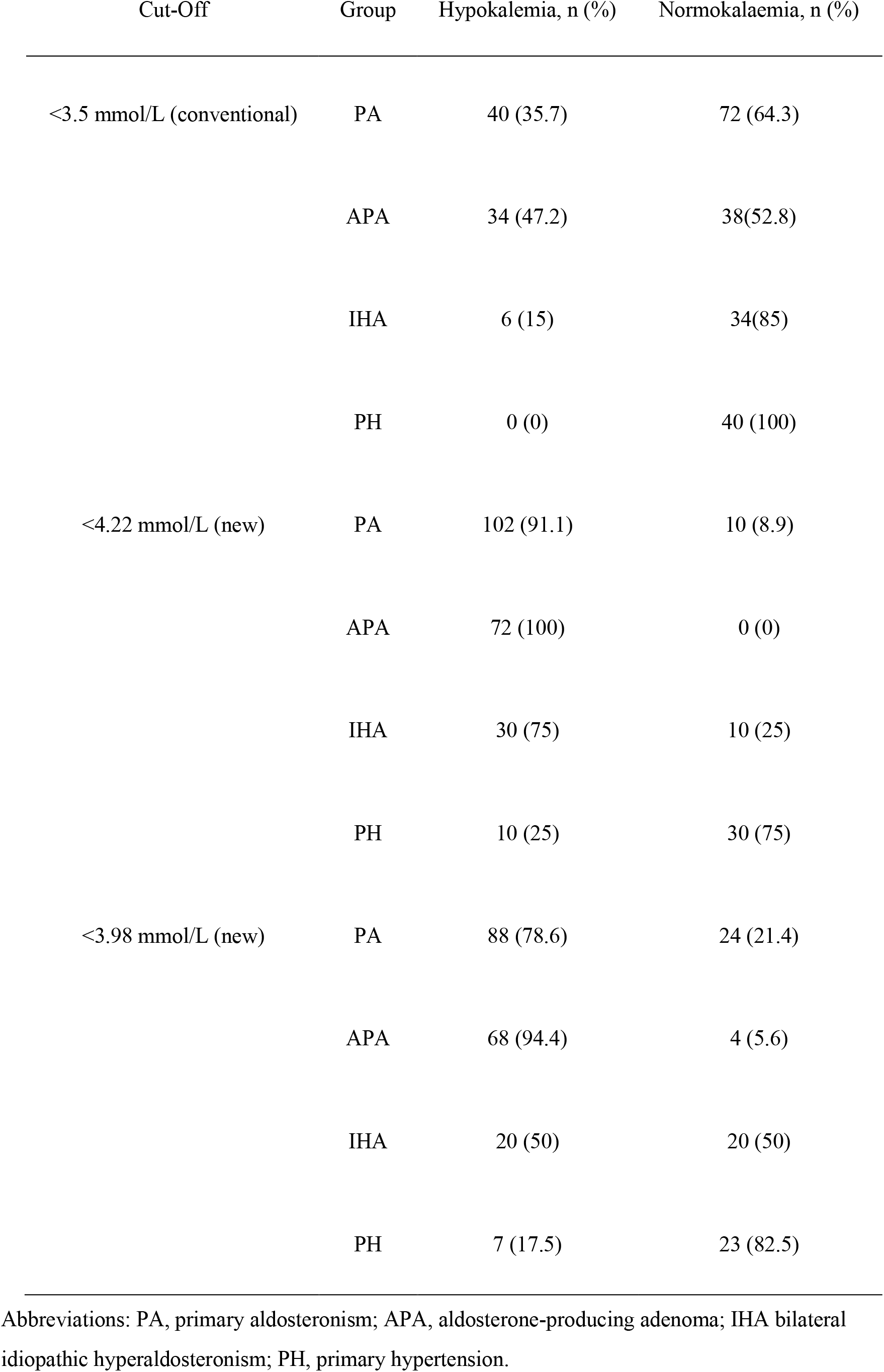
Prevalence of hypokalemia using conventional and new cut-off points.

| Cut-Off | Group | Hypokalemia, n (%) | Normokalaemia, n (%) |
| --- | --- | --- | --- |
| <3.5 mmol/L (conventional) | PA | 40 (35.7) | 72 (64.3) |
|  | APA | 34 (47.2) | 38(52.8) |
|  | IHA | 6 (15) | 34(85) |
|  | PH | 0 (0) | 40 (100) |
| <4.22 mmol/L (new) | PA | 102 (91.1) | 10 (8.9) |
|  | APA | 72 (100) | 0 (0) |
|  | IHA | 30 (75) | 10 (25) |
|  | PH | 10 (25) | 30 (75) |
| <3.98 mmol/L (new) | PA | 88 (78.6) | 24 (21.4) |
|  | APA | 68 (94.4) | 4 (5.6) |
|  | IHA | 20 (50) | 20 (50) |
|  | PH | 7 (17.5) | 23 (82.5) |

Abbreviations: PA, primary aldosteronism; APA, aldosterone-producing adenoma; IHA bilateral idiopathic hyperaldosteronism; PH, primary hypertension.

### 3.5. Combined Diagnostic Strategy

To enhance specificity while retaining improved sensitivity, we evaluated the new potassium cut-off value for patients with ARR>3.7. This combined criterion identified 102 of 112 PA patients (91.1%) while correctly classifying 30 of 40 PH patients (75%) (Table 6). The AUC for the combined criterion was 0.897(95% CI: 0.844–0.949), which was significantly higher than the conventional potassium cut-off (P<0.001).

### 3.6. Subgroup Analysis by PA Subtype

Among APA patients (n=72), the new cut-off of <4.22 mmol/L identified 72 patients (100%), compared with only 34 (47.2%) by the conventional threshold, when the cut-off of <3.98 mmol/L identified 68 patients (94.4%). Among IHA patients (n=40), the new cut-off <4.22 mmol/L identified 30 patients (75%), compared with 6 (15%) by the conventional threshold, when the new cut-off of <3.98 mmol/L identified 20 patients (50%) (Table 6).

## 4. Discussion

This study provides three novel insights. First, we systematically confirm that the conventional hypokalemia threshold identifies fewer than 40% of PA patients. Second, we characterize the left-shifted potassium distribution in PA compared with PH and IHA. Third, and most importantly, we establish an optimized potassium cut-off of 4.22 mmol/L specifically within ARR-positive patients—a population in whom potassium-based refinement has not been previously evaluated.

The observation that only 35.7% of PA patients in our data presented with conventional hypokalemia (<3.5 mmol/L) is consistent with the contemporary literature. Hypokalemia in approximately 30% of PA patients across international centers^7^, and a systematic review reported a prevalence ranging from 9% to 37%^8^. The persistently low prevalence of hypokalemia in PA reflects several factors: dietary potassium intake may partially compensate for renal losses; the severity of aldosterone excess exists on a spectrum; and the widespread use of antihypertensive medications that raise serum potassium (ACE inhibitors, ARBs) may mask the underlying hypokalemic tendency ^9^. Our finding that APA patients had a higher prevalence of hypokalemia (47.2%) than IHA patients (15%) is consistent with the known greater aldosterone secretory capacity of unilateral adenomas^4,6,10^.

The central finding of this study—that the majority of PA patients have serum potassium in the low-normal range—has important clinical implications. The current paradigm of PA screening, particularly in non-specialized settings, often relies on the presence of hypokalemia as a clinical trigger for initiating ARR testing^11,12^. Our data demonstrate that this approach is fundamentally flawed, as it systematically excludes the largest subgroup of PA patients—those with serum potassium between 3.5 and 4.0 mmol/L. In our study, these patients, while not meeting the conventional definition of hypokalemia, still had significantly lower potassium levels than PH patients (mean 3.62 vs. 4.56 mmol/L, P<0.001), reflecting the chronic kaliuretic effect of aldosterone excess. The new serum potassium cut-off of 4.22 mmol/L identified in this study represents a clinically actionable threshold that balances sensitivity and specificity. While the conventional cut-off of 3.5 mmol/L had an unacceptably low sensitivity (35.7%), it was highly specific (100%), reflecting the fact that severe hypokalemia is rare in PH patients.

The new potassium cut-off value for patients with ARR>3.7, further refined diagnostic precision. The new cut-off of 4.22 mmol/L traded a 8.9 percentage point reduction in specificity for a 42 percentage point gain in sensitivity—a favorable trade-off in the context of screening, where the primary goal is to maximize case detection. In clinical practice, for patients already identified as having an elevated ARR (>3.7), applying the new potassium threshold (<4.22 mmol/L) can serve as a secondary filter to identify those with a high probability of confirmed PA. This may help clinicians prioritize patients for confirmatory testing and reduce unnecessary procedures. It should be acknowledged that a specificity of 91.1% implies that approximately 9% of non-PA hypertensive patients would be classified as screen-positive under the new threshold, leading to unnecessary ARR testing and confirmatory procedures. If applied to the general hypertensive population where PA prevalence is approximately 5– 10%, the number of false-positive cases would substantially exceed true-positive cases. Therefore, we recommend that this threshold be used as a clinical alert rather than a definitive diagnostic criterion, and its implementation should be combined with clinical judgment regarding pretest probability. Future studies are needed to evaluate the cost-effectiveness of this approach in real-world settings.

Applying the conventional threshold would have captured only 40 of 112 PA patients in our study, whereas the new threshold alone captured 102. The 155% increase in case detection represents a substantial improvement in screening efficiency, which is particularly relevant given that PA is one of the few curable forms of secondary hypertension. Each missed PA case represents a missed opportunity for targeted therapy—surgical adrenalectomy for APA patients, which offers a 30–60% cure rate for hypertension, or mineralocorticoid receptor antagonist therapy for IHA patients, which provides superior blood pressure and cardioprotective outcomes compared with conventional antihypertensive regimens^13-15^. However, no previous study has systematically determined an optimal potassium cut-off using ROC analysis and the Youden index in a three-group (PA, IHA, PH), nor evaluated the combination of potassium with ARR for enhanced diagnostic precision.

The IHA group in our study provides additional insight into the continuum of aldosterone excess and potassium homeostasis. IHA patients demonstrated an intermediate distribution of serum potassium, with 15% <3.5 mmol/L and 75% <4.22 mmol/L, consistent with their position between PA and PH on the aldosterone excess spectrum. The IHA subgroup merits special attention. With 75% of IHA patients having serum potassium below 4.22 mmol/L, this threshold may identify a significant proportion of patients with subtler aldosterone excess who might otherwise remain undiagnosed under conventional criteria. Given that IHA patients benefit from mineralocorticoid receptor antagonist therapy for blood pressure control and cardioprotection, earlier identification through a relaxed potassium threshold could have therapeutic implications. However, prospective studies are needed to determine whether earlier treatment of these less severe cases translates into improved cardiovascular outcomes.

In most patients, AVS to ascertain the laterality of aldosterone secretory autonomy is the criterion standard, except in young patients (<35 years) with spontaneous hypokalemia and plasma aldosterone concentration >30 ng/dL with a unilateral adenoma >10 mm^16^. We found that serum potassium was lower in APA patients than in IHA patients, and a serum potassium value of 3.98 was used as a cut-off value for differential APA and PH (the specificity was 94.4% and the sensitivity was 82.5%). PA patients with serum potassium lower than 3.98 may be more likely to be diagnosed as APA, which may be helpful for the differential of APA and IHA. This cut-off may improve pre-AVS risk stratification for APA, but it cannot replace AVS for subtype differentiation without independent prospective validation.

Several limitations of this study should be acknowledged. First, the sample size was relatively modest (n=152), and multi-center validation in larger cohorts is needed before the new cut-off can be broadly applied. Second, all serum potassium measurements were performed under standardized conditions after medication washout; in real-world practice, concurrent use of RAAS inhibitors and potassium supplements may affect potassium levels and limit the applicability of the new threshold. Third, this was a cross-sectional study, and we could not evaluate whether earlier identification of PA based on the new cut-off translates into improved cardiovascular outcomes. Fourth, our study was conducted in a Chinese population at a single tertiary center; the optimal potassium cut-off may differ across ethnic groups and practice settings due to variations in dietary potassium intake, genetic predisposition, and assay methodology. Fifth, serum potassium is subject to biological variability, and a single measurement may not adequately represent a patient’s chronic potassium status.

In conclusion, this study demonstrates that the conventional serum potassium cut-off of 3.5 mmol/L is an insensitive screening criterion for PA, missing over 60% of confirmed cases. PA patients have a characteristic leftward shift in serum potassium distribution, with the majority residing in the low-normal range (3.5–4.0mmol/L). A new serum potassium cut-off of 4.22mmol/L significantly improves screening sensitivity and when combined with ARR>3.7, significantly improves diagnostic accuracy. We propose that hypertensive patients with serum potassium <4.22 mmol/L should be prioritized for PA-specific screening, regardless of whether they meet the conventional definition of hypokalemia.

This simple, accessible, and cost-effective strategy may substantially improve PA case detection and facilitate timely intervention for this treatable cause of secondary hypertension.

## Data Availability

All data covered in the manuscript are available

